# Association Between Abdominal Obesity, Cardiovascular Fitness, and Incident Hypertension in Middle-Aged Korean Adults

**DOI:** 10.1101/2024.12.13.24319025

**Authors:** Jisuk Chae, Mihee Kim, Soo young Jung, Seung Don Yoo, Si-Hyuck Kang, Kyesan Lee, Jung-Hyun Kim, Junga Lee

## Abstract

**BACKGROUND:** Hypertension is a global health crisis that significantly increases the risk of cardiovascular diseases, stroke, and chronic kidney disease. Despite advances in treatment, the prevalence of hypertension continues to rise. It is important to know the association between abdominal obesity, cardiovascular fitness, and hypertension in middle-aged Koreans to develop effective prevention and management strategies for hypertension. This study aims to investigate the association between body composition, physical fitness, and hypertension in middle-aged Korean population. We examined the impact of abdominal obesity and cardiovascular fitness on the risk of hypertension.

**METHODS and RESULTS:** A cross-sectional study was conducted among 60 healthy adults (mean aged 54.23±7.34) recruited. Participants underwent assessments of anthropometry, body composition, blood pressure, and physical fitness. The primary outcome was the prevalence of hypertension. Secondary outcomes included body composition measures (body fat mass, body lean mass and waist circumference), and physical fitness assessments (grip strength, sit-ups, sit-and-reach test, and YMCA step test). Participants with hypertension had significantly higher body fat mass and waist circumference compared to those without hypertension. The risk of hypertension was significantly increased by 17.7% with a 1 kg increase in body fat and by 11.9% with a 1 cm increase in waist circumference. The risk of hypertension was significantly increased in the group with hypertension accompanied by abdominal obesity as cardiovascular fitness, measured by the YMCA step test, decreased (Adjusted OR: 0.326, 95% CI: 0.168-0.631).

**CONCLUSION:** These findings highlight the significant association between abdominal obesity, reduced cardiovascular fitness, and hypertension in middle-aged Korean adults. Abdominal obesity and low cardiovascular fitness were identified as independent risk factors for hypertension in this population. These results suggest that weight management to reduce abdominal obesity and regular physical activity to promote cardiovascular fitness are key to the prevention and management of hypertension.

## Introduction

Hypertension is a significant global health problem that can lead to serious health complications, such as heart disease, stroke, and kidney disease^1–3^. The number of people with hypertension worldwide was more than doubled from 650 million in 1990 to 1.27 billion in 2019 by the World Health Organization, which emphasized the increasing severity of this health issue^4^. Middle-aged individuals are considered to be at a higher risk of hypertension due to lifestyle and hormonal changes^5^. The risk and mortality of cardiovascular disease in older age are significantly increased by inadequate management of high blood pressure in middle-aged^5^. A global prevalence of 18.6 million cases of hypertensive heart disease was revealed by research published in 2024^6^. Young adults with prehypertension (systolic blood pressure (SBP): 120-129 mmHg, diastolic blood pressure (DBP): 80-84 mmHg) were found to be 1.19 times more prone to experiencing cardiovascular events compared to those with normal blood pressure (SBP <120 mmHg, DBP <80 mmHg) by a recent study that examined the relationship between hypertension and cardiovascular disease^7^. A recent study from 2024 revealed that the prevalence of chronic kidney disease caused by hypertension increased by 161.97% compared to the 1990s, with over 1.57 million cases reported in 2019 alone^8^. Hypertension has been linked to brain blood vessel damage by multiple studies, which raises the probability of both stroke and Alzheimer’s disease^9–11^ A significant positive association between hypertension and stroke was revealed by a decade-long study of U.S. adults ^12^, with hypertensive individuals found to be 3.9 times more prone to experiencing hemorrhagic stroke^13^. A 2019 meta-analysis found that middle-aged hypertension was associated with a 25% increased risk of Alzheimer’s disease^14^. Moreover, untreated hypertension was identified as an independent risk factor for Alzheimer’s disease when compared to controlled hypertension or a healthy control group^15^.

The importance of cardiovascular fitness in the control of hypertension has been emphasized by recent studies. Cardiovascular fitness has been shown to have a stronger relation to hypertension than physical activity^16^. An increased risk of hypertension was found in individuals with low cardiovascular fitness^16,17^. This relationship can be explained by the correlation between aerobic exercise and cardiovascular fitness. Aerobic exercise has been shown to be the most effective way to improve cardiovascular fitness^18,19^, and has a positive impact on hypertension management. Regular aerobic exercise has been shown to increase cardiac output as the heart’s function is enhanced, the heart’s workload is decreased, and blood pressure is reduced^20,21^. It has also been shown that blood pressure is lowered by a reduction in vascular resistance and the inhibition of the activity of the sympathetic nervous system and the renin-angiotensin-aldosterone system^22^. Lower levels of physical activity have been associated with higher blood pressure, which is a major risk factor for cardiovascular disease^23,24^. Weight gain and increased body fat can be caused by decreased physical fitness and lack of exercise, which can lead to obesity and impaired blood pressure regulation, and that can lead to hypertension^25^.

Waist circumference and abdominal obesity have been strongly associated with the development of hypertension^26–30^. High waist circumference has been shown to be a strong predictor of visceral adiposity, rather than a simple measure of overall body weight^31^. Visceral fat has been shown to not only store energy but also secrete various bioactive substances that increase the risk of metabolic diseases, cardiovascular diseases, and other chronic conditions^32,33^. Abdominal obesity is strongly associated with metabolic syndrome, including hypertension, diabetes, and dyslipidemia, and is a significant risk factor for heart disease^34^. Meta-analysis has confirmed that abdominal obesity is a strong predictor of hypertension^35^. As waist-to-height ratio (WHtR) has increased, systolic and diastolic aortic blood pressure, systemic vascular resistance, and pulse wave velocity have also been shown to increase^36^. Visceral fat accumulation has been shown to have a strong correlation with hypertension^37^. Excessive nutrient intake has been shown to lead to enlarged visceral fat, which over time has been shown to damage tissues through endocrine stress, inducing inflammatory responses^38,39^, and disrupting glucose and lipid metabolism, thereby contributing to the development of hypertension^39,40^. Therefore, weight loss and reduction in body fat are indispensable components of hypertension management^41^.

This study investigated the associations between numerous variables, including age, sex, height, body composition, and physical fitness levels, and the prevalence of hypertension among middle-aged adults. The relationship among abdominal obesity and cardiovascular fitness on the onset of hypertension was examined.

## Methods

### 1. Study Participants

Sixty healthy middle-aged adults were recruited for this study. Inclusion criteria were as follows: (1) middle-aged Korean adults and (2) no history of systematic exercise in the past six months. Exclusion criteria included: (1) pregnancy and (2) pre-existing musculoskeletal, cardiovascular, or immunological conditions that could impair exercise participation. Participants were recruited via poster advertisements displayed in the laboratory.

This study was approved by the Institutional Review Board (IRB File No. KHGIRB-23-468) and conducted in accordance with ethical guidelines.

### 2. Experimental Methods and Procedures

#### 2.1. Pre-experiment Preparation

Participants visited the laboratory once. One day prior to their visit, participants were instructed via phone to refrain from consuming food, alcohol, and caffeine for at least 8 hours before the visit, and to avoid strenuous exercise for 24 hours preceding the visit. After arriving, participants were informed about the study, provided consent, and were checked to ensure they met the study’s requirements. Anthropometric measurements, body composition, physical fitness assessment and resting metabolic rate (RMR) were measured for all participants before the exercise. All measurements were performed using the same method by experienced researchers.

#### 2.2. Anthropometric Measurements and Body Composition

Participants were instructed to wear comfortable clothing and athletic shoes for the physical fitness assessment. Anthropometric measurements, including height and weight, were obtained using a stadiometer (GL-150Tech, G-Tech International, Korea). Body mass index (BMI) was calculated as weight (kg) divided by height squared (m²). Waist and hip circumferences were measured using a tape measure, and the waist-to-hip ratio was calculated.

Blood pressure and heart rate were measured using an automated blood pressure monitor (JPN710T, OMRON, China). Participants were seated and allowed to rest for at least 10 minutes prior to measurement. The blood pressure cuff was positioned 2 cm above the antecubital crease of the left arm, at heart level. Systolic and diastolic blood pressures were recorded.

Body composition, including body fat percentage and fat mass, were obtained using a bioelectrical impedance analysis device (Inbody 270, Inbody, Korea).

#### 2.3. Physical Fitness Assessment

##### (1) Upper Body Strength

A dynamometer (TKK5401, TAKEI, Japan) was used to evaluate grip strength, which is an indicator of upper body strength. Participants assumed a standing posture with feet shoulder-width apart and grasped the handle with their dominant hand, ensuring a comfortable grip with the second joint of the fingers. The device’s indicator was positioned facing outward. With arms fully extended and the torso inclined at a 15° angle, participants exerted maximum force on the handle for 3 seconds. This procedure was repeated twice for each hand, and the highest value was recorded to the nearest 0.1 kg. Relative grip strength was calculated using the following formula:

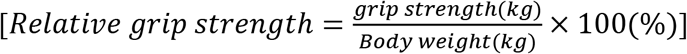

##### (2) Muscular Endurance

Sit-ups were measured as an indicator of muscular endurance. Participants lay on a mat with their head and back flat, knees bent, feet flat on the floor, and feet hip-width apart. They placed their hands on their thighs with arms extended and performed sit-ups, touching the fingertips of the assessor who was holding their knees. The maximum number of repetitions in 1 minute was recorded.

##### (3) Flexibility

The sit-and-reach test was used to assess flexibility. Participants sat on the floor with their legs extended and feet against a sit-and-reach box. They placed one hand on top of the other and reached forward as far as possible, holding the position for 2 seconds. This was repeated twice, and the highest value was recorded. Participants were instructed to avoid using momentum, bending their knees, or lifting their feet from the box.

##### (4) Cardiovascular Fitness

The YMCA step test was used to assess cardiovascular fitness. Participants stepped up and down on a 30.5 cm step box at a rate of 96 beats per minute for 3 minutes. After 3 minutes, trained sports medicine researchers measured participants’ heart rate in a seated position. Heart rate was recorded from the participants’ radial artery for 1 minute. Maximal oxygen consumption was estimated using the following formula: This equation predicts a male and female’s maximal oxygen consumption based on their age, height, weight, and heart rate recovery after one minute of exercise.

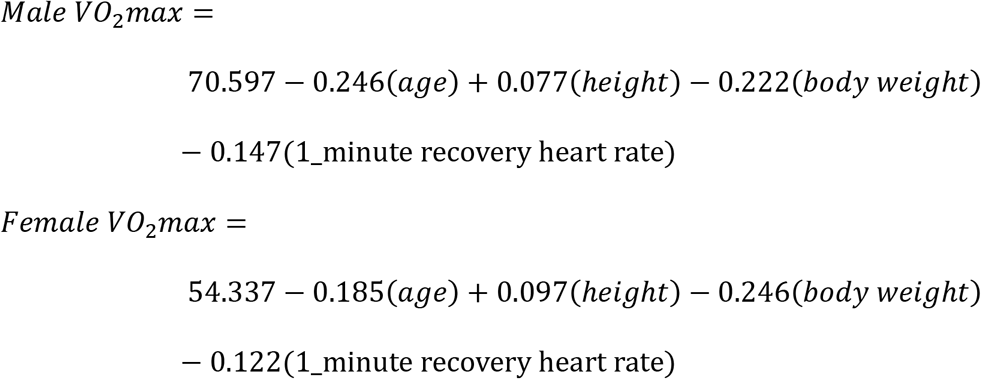

#### 2.4 Resting Metabolic Rate Measurement

Before measuring resting metabolic rate (RMR), participants were asked to fast for 8 hours and to avoid strenuous physical activity on the day of the measurement. Participants rested seated for 20 minutes before testing at the laboratory. A mask was fitted, and oxygen consumption and carbon dioxide production were measured for 15 minutes using a wireless gas analyzer (K5, Cosmed, Italy).

### 3. Data Analysis

Data were analyzed using SPSS version 25.0. A statistical significance level of p < 0.05 was adopted throughout the study. The general characteristics of the participants were analyzed using descriptive statistics and frequency analysis.

Independent t-tests and chi-square or Fisher’s exact tests were used to compare clinical characteristics between middle-aged Korean adults with and without abdominal obesity, as appropriate.

The association between hypertension diagnosis and body composition or abdominal obesity was analyzed using binary logistic regression analysis. This statistical method is employed when the dependent variable is presence of hypertension and the aim is to assess the influence of independent variables (body fat mass and skeletal muscle mass) on the dependent variable.

The relationships between blood pressure, abdominal obesity, and physical fitness assessments (grip strength, sit-ups, sit-and-reach test, and YMCA step test) were analyzed using multinomial logistic regression. The dependent variables were blood pressure (normal blood pressure, prehypertension, hypertension) and abdominal obesity (normal waist circumference, abdominal obesity). The independent variables included measures of physical function and body composition.

By comparing the β coefficients and p-values across different groups, we set the significance level at 0.05.

## Results

The general characteristics and physical fitness levels of the 60 middle-aged Korean adults who participated in this study are presented in Table 1. The average age was found to be similar for both genders, around 54 years old. Men were found to have an average height of 1.73m, approximately 13cm taller than women (1.60m). Men were found to weigh an average of 74.56kg, about 16kg heavier than women (58.31kg). Body mass index (BMI), an indicator of obesity, was found to be slightly higher in men (24.98 kg/m²) compared to women (22.75 kg/m²). Although men and women were found to have similar average body fat mass (17.73kg for men, 18.47kg for women), men were found to have a lower body fat percentage (22.93%) compared to women (30.71%). Men were found to have a larger waist circumference (92.09cm) compared to women (85.41cm), with a difference of about 7cm. The waist-to-hip ratio (WHR) was found to be slightly higher in men (0.92) than in women (0.88). Heart rate was found to be similar between genders. Systolic blood pressure was found to be approximately 8mmHg higher in men (130.40mmHg) compared to women (122.70mmHg). Diastolic blood pressure was also found to be higher in men (85.37mmHg) compared to women (76.87mmHg), with a difference of about 9mmHg. Grip strength was found to be significantly higher in men (40.70kg) compared to women (25.24kg). Men were found to perform an average of 25.47 sit-ups, approximately 6 more repetitions than women (19.93). The step test results, indicating cardiovascular fitness, showed that men had a higher average value (27.80 ml/kg/min) compared to women (19.79 ml/kg/min), with a difference of about 8 ml/kg/min. Men were found to have a shorter sit-and-reach distance (6.63cm) compared to women (14.74cm), with a difference of about 8cm.

**Table 1.** Clinical characteristics among the 40-69-year-old Korean adults.

|  | *n*=60 | Male (*n*=30) | Female (*n*=30) |
| --- | --- | --- | --- |
| Age (years) | 54.23±7.34 | 54.40±6.99 | 54.07±7.79 |
| Male/Female, *n* (%) | 30 (50) / 30 (50) | - | - |
| Height (m) | 1.66±0.08 | 1.73±0.06 | 1.60±0.04 |
| Weight (kg) | 66.44±14.27 | 74.56±13.02 | 58.31±10.40 |
| BMI (kg/m<sup>2</sup>) | 23.86±3.99 | 24.98±3.79 | 22.75±3.94 |
| Body Lean Mass (kg) | 26.79±6.22 | 32.00±4.08 | 21.59±2.48 |
| Body Fat Mass (kg) | 18.10±7.48 | 17.73±7.39 | 18.47±7.67 |
| Body Fat Percentage (%) | 26.82±7.63 | 22.93±6.30 | 30.71±6.88 |
| Waist Circumference (cm) | 88.75±10.84 | 92.09±9.91 | 85.41±10.85 |
| WHR | 0.90±0.06 | 0.92±0.05 | 0.88±0.05 |
| Heart Rate (bpm) | 70.70±9.23 | 70.80±8.88 | 70.60±9.72 |
| SBP (mmHg) | 126.55±16.45 | 130.40±13.92 | 122.70±18.06 |
| DBP (mmHg) | 81.12±10.69 | 85.37±8.70 | 76.87±10.92 |
| Physical Fitness Components |  |  |  |
| Hand grip (kg) | 32.97±9.92 | 40.70±7.83 | 25.24±3.95 |
| Sit-ups (reps) | 22.70±11.24 | 25.47±11.83 | 19.93±10.06 |
| Step test (ml/kg/min) | 23.80±5.50 | 27.80±4.20 | 19.79±3.27 |
| Sit and reach test (cm) | 10.69±8.56 | 6.63±7.94 | 14.74±7.21 |
| RMR (Kcal) | 1980.3±394.38 | 2188.73±353.36 | 1772.03±319.00 |

Mean±SD. *p*-values of the continuous variables were compared using independent samples *t*-tests. BMI, body mass index; DBP, diastolic blood pressure; RMR, resting metabolic rate; SBP, systolic blood pressure; WHR, waist to hip ratio

The associations between hypertension diagnosis and body composition were examined in a binary logistic regression analysis, the results of which are presented in Table 2. A significant positive association was found between body fat mass and the risk of hypertension (Crude Odds Ratio (OR): 1.141, 95% CI: 1.029-1.266, Adjusted OR: 1.148, 95% CI: 1.017-1.296, p<0.05), suggesting that individuals with greater body fat mass were at a higher risk of developing hypertension. However, no statistically significant association was found between skeletal muscle mass and hypertension diagnosis.

**Table 2.** Association between Hypertension Diagnosis and Body Composition.

| Group | Variables | Crude ORs<br>(95% CI) | Adjusted ORs<br>(95% CI) |
| --- | --- | --- | --- |
| Normal<br>(BP < 130/80 mmHg) | Body Fat mass (kg) | 1 (reference) | 1 (reference) |
|  | Skeletal muscle mass (kg) |  |  |
| Hypertension<br>(130/80 mmHg ≤ BP) | Body Fat mass (kg) | 1.141(1.029-1.266) * | 1.148(1.017-1.296) * |
|  | Skeletal muscle mass (kg) | 1.042(0.933-1.165) | 1.177(0.907-1.528) |

Binary logistic regression analysis. Adjusted ORs are ORs adjusted for the confounding factor such as age and gender. \**p*<0.05, †*p*<0.01, and ‡*p*<0.001. BP, blood pressure.

The results presented in Table 3 indicate that a significant positive association was found between waist circumference and the risk of hypertension. Binary logistic regression analysis demonstrated that a 1 cm increase in waist circumference was associated with a 10% increased odds of hypertension diagnosis. Even after adjusting for age and sex, this association remained significant. The result suggests that abdominal obesity was found to be an independent risk factor for hypertension.

**Table 3.** Association between Hypertension Diagnosis and Abdominal Obesity.

| Variables | Group | Crude ORs (95% CI) | Adjusted ORs (95% CI) |
| --- | --- | --- | --- |
| Waist Circumference (cm) | Normal (BP < 130/80 mmHg) | 1 (reference) | 1 (reference) |
|  | Hypertension (130/80 mmHg ≤ BP) | 1.097(1.028-1.170)<sup>†</sup> | 1.119(1.037-1.208)<sup>†</sup> |

Binary logistic regression analysis. Adjusted ORs are ORs adjusted for the confounding factor such as age and gender. \* $p < 0.05$ , <sup>†</sup> $p < 0.01$ , and <sup>‡</sup> $p < 0.001$ . BP, blood pressure.

The relationship between hypertension, abdominal obesity, and physical fitness components was examined using a multinomial logistic regression analysis, the results of which are presented in Table 4. These findings indicated that the relationship between physical fitness and hypertension was influenced by the presence of abdominal obesity. Among individuals without abdominal obesity, no significant associations were found between grip strength, sit-ups, step test, sit-and-reach test, and the risk of developing hypertension. However, among those with abdominal obesity, individuals in the prehypertension stage with lower step test scores were found to have a significantly increased risk of developing hypertension (Crude OR: 0.849, 95% CI: 0.655-1.102, Adjusted OR: 0.455, 95% CI: 0.259-0.801). Similarly, in the hypertension group, those with lower step test scores were found to have a significantly increased risk of hypertension (Crude OR: 0.670, 95% CI: 0.490-0.917, Adjusted OR: 0.326, 95% CI: 0.168-0.631). While no significant association was found between physical fitness and hypertension risk in those without abdominal obesity, this study results suggested that lower step test scores were significantly associated with a higher risk of hypertension in individuals with abdominal obesity. This implied that abdominal obesity strengthened the link between aerobic capacity and the risk of developing hypertension.

**Table 4.** Association between Hypertension, Abdominal Obesity, and Physical Fitness Components (grip strength, sit-ups, step test, and sit-and-reach test)

| Group | Variables | Crude ORs (95% CI) | Adjusted ORs (95% CI) |
| --- | --- | --- | --- |
| Normal BP & Normal WC | Hand grip (kg) | 1 | 1 |
|  | Sit-ups (reps) |  |  |
|  | Step test (ml/kg/min) |  |  |
|  | Sit and reach test (cm) |  |  |
| Normal BP & Abdominal Obesity | Hand grip (kg) | 1.097(0.959-1.256) | 1.048(0.826-1.328) |
|  | Sit-ups (reps) | 0.990(0.889-1.102) | 0.983(0.872-1.108) |
|  | Step test (ml/kg/min) | 0.777(0.594-1.017) | 0.672(0.425-1.064) |
|  | Sit and reach test (cm) | 0.968(0.846-1.108) | 0.975(0.838-1.136) |
| Prehypertension & Normal WC | Hand grip (kg) | 1.030(0.917-1.156) | 0.984(0.812-1.193) |
|  | Sit-ups (reps) | 0.965(0.875-1.065) | 0.971(0.880-1.072) |
|  | Step test (ml/kg/min) | 1.096(0.887-1.352) | 0.996(0.679-1.460) |
|  | Sit and reach test (cm) | 1.017(0.901-1.148) | 1.024(0.903-1.161) |
| Prehypertension & Abdominal Obesity | Hand grip (kg) | 1.137(0.994-1.299) | 0.878(0.699-1.103) |
|  | Sit-ups (reps) | 0.958(0.861-1.066) | 1.003(0.885-1.137) |
|  | Step test (ml/kg/min) | 0.849(0.655-1.102) | 0.455(0.259-0.801)<sup>†</sup> |
|  | Sit and reach test (cm) | 0.957(0.836-1.096) | 0.992(0.846-1.164) |
| Hypertension & Normal WC | Hand grip (kg) | 1.113(0.903-1.373) | 0.900(0.640-1.265) |
|  | Sit-ups (reps) | 0.921(0.783-1.082) | 0.942(0.784-1.132) |
|  | Step test (ml/kg/min) | 0.852(0.570-1.274) | 0.525(0.241-1.145) |
|  | Sit and reach test (cm) | 1.067(0.859-1.327) | 1.104(0.862-1.414) |
| Hypertension & Abdominal Obesity | Hand grip (kg) | 1.224(1.051-1.426)<sup>†</sup> | 0.893(0.686-1.163) |
|  | Sit-ups (reps) | 0.910(0.802-1.033) | 0.947(0.841-1.102) |
|  | Step test (ml/kg/min) | 0.670(0.490-0.917)<sup>*</sup> | 0.326(0.168-0.631)<sup>‡</sup> |
|  | Sit and reach test (cm) | 0.939(0.804-1.097) | 1.013(0.844-1.216) |

Multinomial logistic regression analysis. Adjusted ORs are ORs adjusted for the confounding factor such as age and gender. \* $p < 0.05$ , <sup>†</sup> $p < 0.01$ , and <sup>‡</sup> $p < 0.001$ . BP, blood pressure; WC, waist circumference.

## Discussion

The relationship between increased body fat mass, abdominal obesity, low cardiovascular fitness, and hypertension in middle-aged adults was investigated in this study. Hypertensive patients had significantly higher body fat mass than patients with normal blood pressure, and the risk of hypertension increased further in individuals with abdominal obesity. A higher risk of hypertension was observed in those with low cardiovascular fitness. These results suggested that increased body fat mass, abdominal obesity, and low levels of physical fitness were significant risk factors for the development of hypertension.

This study was found to have a significantly increased risk of hypertension with increasing body fat mass and waist circumference. After adjusting for sex and age, each 1 kg increase in body fat was associated with a 14.8% higher risk of developing hypertension (Odds Ratio (OR) 1.148; 95% CI 1.017-1.296), and each 1 cm increase in waist circumference was associated with an 11.9% higher risk. Abdominal obesity was identified as one of the strongest risk factors for hypertension. A recent study involving 4,658 diabetic patients in seven communities in Shanghai, China, in 2018 revealed a significant positive correlation between abdominal obesity and the prevalence of cardiovascular disease and diabetic kidney disease. Among men, each one-standard deviation increase in visceral fat was associated with 1.35-fold higher odds of cardiovascular disease (95% CI: 1.13-1.62) and 1.38-fold higher odds of diabetic kidney disease (95% CI: 1.12-1.70). For women, a similar increase in visceral fat was linked to 1.32-fold higher odds of cardiovascular disease (95% CI: 1.04-1.69) and 2.50-fold higher odds of diabetic kidney disease (95% CI: 1.81-3.47)^42^. Visceral fat accumulation, which is caused by abdominal obesity, increases insulin resistance, increases blood glucose concentration, strains the heart, reduces heart function, and hardens blood vessels to impede blood circulation^43^. A recent study that investigated the association between waist circumference and the prevalence of hypertension in 27,894 American adults in 2023^26^ and an analysis of 27 papers (18 cross-sectional studies and 9 cohort studies) published between 2010 and 2023 reported that high abdominal visceral fat was related to the development of cardiovascular disease^32^. Inflammatory substances released by abdominal fat harm vascular endothelium, which led to increased atherosclerosis, impaired blood pressure regulation, and heightened heart strain, thus this could lead to heart failure^38^. The heart is made to work harder to pump blood around the body by excess body fat, which can cause the heart muscle to thicken over time^44,45^. Cardiac hypertrophy, a thickened heart muscle is pumped harder, which raises blood pressure and puts more strain on blood vessels. This can contribute to the development of cardiovascular diseases, such as arteriosclerosis^44,46^. In other words, the accumulation of visceral fat associated with abdominal obesity could increase the risk of various cardiovascular diseases, with hypertension, atherosclerosis, coronary artery disease, cerebrovascular disease, and heart failure^32^. A recent study investigating the correlation between abdominal obesity and insulin resistance has revealed that visceral fat cells secrete inflammatory cytokines that contribute to insulin resistance^42,47^. Abdominal obesity could cause visceral fat accumulation and promote the secretion of inflammatory cytokines, which could impair vascular endothelial function and increase blood pressure^48,49^. A comprehensive review of research published before October 2022 indicated that adipokines, hormones produced by fat cells, play a central role in the inflammation associated with obesity. These hormones release substances that harm the inner lining of blood vessels, increasing the risk of conditions such as hardening of the arteries, high blood pressure, and heart disease^48^. Also, adipose tissue contributed to elevated blood pressure by activating the renin-angiotensin-aldosterone system^50,51^. These findings are consistent with the proposed mechanism whereby visceral fat accumulation triggers insulin resistance, promotes the release of inflammatory cytokines, and activates the renin-angiotensin-aldosterone system, ultimately resulting in endothelial dysfunction and hypertension^52,53^.

This study was found to have individuals with low cardiovascular fitness at a higher risk of developing hypertension. After adjusting for age and sex, the odds ratio (OR) was 0.326 (95% CI, 0.168-0.631), indicating that the group with both hypertension and abdominal obesity had the lowest cardiovascular fitness. This suggested a direct link between decreased cardiovascular fitness and impaired blood pressure control. Several mechanisms contributed to the association between low cardiovascular fitness and an increased risk of hypertension. In a recent study of 138 overweight or obese adults, cardiovascular fitness was found to be associated with insulin sensitivity. Lower cardiovascular fitness was linked to reduced insulin sensitivity, which increased the risk of cardiovascular disease and type 2 diabetes^54^. Low cardiovascular fitness can make it difficult for sufficient oxygen to be supplied during physical activity, which can cause tissue damage and chronic inflammation. This, in turn, can worsen vascular damage and lead to increased blood pressure^55^. Low cardiovascular fitness can cause decreased vascular endothelial function, which can reduce the ability of blood vessels to expand, making it difficult to control blood pressure^56^ It can also weaken the pumping function of the heart, leading to poor blood circulation and increased blood pressure^57^. Excessive activation of the sympathetic nervous system due to a lack of physical activity can cause vasoconstriction and increase blood pressure^58^. On the other hand, regular exercise has the effect of lowering blood pressure through the reduction of body fat, increased insulin sensitivity, and improved vascular endothelial function^59^.

However, this study found no significant association between muscular strength and flexibility (measured by grip strength, sit-ups, and the sit-and-reach test) and hypertension. While previous research suggested a potential positive influence of muscular strength on blood pressure control^60^, further investigation is necessary.

A strong correlation has been found between abdominal obesity, hypertension, and cardiovascular health. The range of motion of the diaphragm is restricted by abdominal obesity, which led to a decrease in lung capacity. This affected cardiac output and blood oxygen saturation, and was considered a major factor that reduced cardiopulmonary function^61^. Low cardiovascular fitness is associated with excessive activation of the sympathetic nervous system, decreased vasodilation capacity, and chronic inflammatory conditions^57,62^. which make blood pressure control difficult and negatively affect the management and treatment of hypertension^63^. This study shows that various mechanisms may act in combination to reduce cardiopulmonary fitness in patients with hypertension accompanied by abdominal obesity. Abdominal obesity and hypertension are the main causes of decreased cardiopulmonary function. Therefore, in order to prevent and manage hypertension and abdominal obesity, it is important to improve cardiovascular fitness through regular aerobic exercise and healthy eating habits. It is understood that abdominal obesity, high blood pressure, and cardiovascular fitness interact with each other and are important factors affecting health. The relationship between these three factors must be understood, and efforts should be made to maintain a healthy life through proper management. This study revealed a relationship between the occurrence of hypertension and various factors, and several strengths were confirmed. First, rather than blood pressure alone being examined, a wide range of physical factors were explored to understand the complex etiology of hypertension.

Second, after age and sex were controlled for, a strong correlation between hypertension and other variables was confirmed. Lastly, this study indicated by the research that enhancing cardiorespiratory endurance could potentially be used to prevent and manage high blood pressure in clinical settings. Despite its strengths, this study was limited by its small sample size, drawn from a specific population of middle-aged Korean adults. The cross-sectional design also limited the ability to establish causality. Furthermore, potential confounders, including dietary and exercise habits, were not considered, nor were genetic factors, which limited the interpretation of the results. Thus, although an association between hypertension, abdominal obesity, body composition, and physical fitness was suggested, the results were limited by factors such as the nonrepresentative sample size, cross-sectional design, and lack of consideration of potential confounders and genetic factors.

The purpose of this study was to examine the association between body fat mass, abdominal obesity, and cardiovascular fitness with the incidence of hypertension in middle-aged adults. This study found that increased body fat mass and abdominal obesity were significant risk factors for hypertension. The risk of decreased cardiovascular fitness is further increased when hypertension is accompanied by abdominal obesity. Decreased cardiovascular fitness and hypertension were mediated by abdominal obesity, which in turn reinforced their synergistic effect.

These findings have important clinical implications for the management of hypertensive patients. First, lifestyle modifications, including weight loss and reductions in abdominal obesity, should be encouraged by healthcare providers for hypertensive patients. Second, personalized exercise prescriptions targeting cardiovascular fitness improvement through regular aerobic exercise, a low-calorie diet for fat reduction, and resistance training to increase basal metabolic rate can enhance the effectiveness of hypertension management.

In conclusion, this study demonstrated that weight loss, including reduced abdominal obesity, and enhanced cardiovascular fitness were essential for the prevention and control of hypertension in middle-aged individuals. Therefore, healthcare providers should encourage lifestyle modifications and exercise among hypertensive patients to prevent complications.

## Data Availability

The datasets used and/or analysed during the current study available from the corresponding author on reasonable request. The datasets generated and/or analysed during the current study are available from the corresponding authors upon reasonable request.

## Acknowledgments

The completion of this research paper would not have been possible without the support and guidance of Junga Lee, Sports Medicine and Science Lab. Her dedication and overwhelming attitude towards helping her students is solely responsible for completing my research paper. The encouragement and insightful feedback were instrumental in accomplishing this task.

Special thanks to Mihee Kim and Soo young Jung for data collection, coding and clinical research assistance. Their extraordinary contributions have greatly improved the quality of this thesis paper.

